# Extracting the Sample Size From Randomized Controlled Trials in Explainable Fashion Using Natural Language Processing

**DOI:** 10.1101/2024.07.09.24310155

**Authors:** Paul Windisch, Fabio Dennstädt, Carole Koechli, Robert Förster, Christina Schröder, Daniel M. Aebersold, Daniel R. Zwahlen

**Affiliations:** Department of Radiation Oncology, Cantonal Hospital Winterthur, Winterthur, Switzerland; Department of Radiation Oncology, Inselspital, Bern University Hospital, University of Bern, Bern, Switzerland

## Abstract

**Background:** Extracting the sample size from randomized controlled trials (RCTs) remains a challenge to developing better search functionalities or automating systematic reviews. Most current approaches rely on the sample size being explicitly mentioned in the abstract.

**Methods:** 847 RCTs from high-impact medical journals were tagged with six different entities that could indicate the sample size. A named entity recognition (NER) model was trained to extract the entities and then deployed on a test set of 150 RCTs. The entities’ performance in predicting the actual number of trial participants who were randomized was assessed and possible combinations of the entities were evaluated to create predictive models.

**Results:** The most accurate model could make predictions for 64.7% of trials in the test set, and the resulting predictions were within 10% of the ground truth in 96.9% of cases. A less strict model could make a prediction for 96.0% of trials, and its predictions were within 10% of the ground truth in 88.2% of cases.

**Conclusion:** Training a named entity recognition model to predict the sample size from randomized controlled trials is feasible, not only if the sample size is explicitly mentioned but also if the sample size can be calculated, e.g., by adding up the number of patients in each arm.

## Introduction

Using natural language processing (NLP) for data mining in biomedical research has been a topic of longstanding interest.^1,2^ Recently, it has gained new interest due to the emerging capabilities of large language models.^3^ Furthermore, new technologies that enable more robust data extraction have been developed in recent years.^4,5^

In clinical research, randomized controlled trials (RCTs) are the gold standard for testing the effectiveness of an intervention.^6^ Therefore, they are the focus of many meta-research efforts, such as systematic reviews or meta-analyses. Automatically extracting PICO (patient, intervention, control, outcome) characteristics from RCTs using NLP could improve various processes, from screening trials over assessing adherence to reporting standards to ultimately fully automating the process of evidence synthesis.^7,8^

A key characteristic of an RCT is the sample size, i.e., the number of people included in the trial.^9^ This information is normally already presented in the abstract, which makes it a suitable parameter for data mining based on the abstract. From a practical point of view, the abstract is also usually not locked behind a paywall.

Inclusion in the trial is usually defined as having undergone randomization.^5^ However, there are different ways to present this information in an RCT. While some trials might explicitly state the number of participants that were randomized, others might just state the number of patients who “were included”, “were analyzed”, or “completed the trial.”

We hypothesized that each of these different phrases carries a different likelihood of the number being presented actually representing the number of patients who were randomized. We, therefore, trained a named entity recognition model to extract these phrases as different entities and build a prediction model that considers the different performances for predicting the ground truth of how many people underwent randomization.

## Methods

A random sample of 996 randomized controlled trials (RCTs) from seven major journals (British Medical Journal, JAMA, JAMA Oncology, Journal of Clinical Oncology, Lancet, Lancet Oncology, New England Journal of Medicine) published between 2010 and 2022 were labeled. To do so, abstracts were retrieved as a txt file from PubMed and parsed using regular expressions (i.e., expressions that match certain patterns in text). For each trial, the number of people who were randomized was retrieved by looking at the abstract, followed by the full publication if the number could not be determined with certainty from the abstract.

In addition, six different entities were tagged in each abstract, independent of whether the information was presented using words or integers. If the number of people who were randomized was explicitly stated (e.g., using the words “randomly,” “randomized,” etc.), this was tagged as “RANDOMIZED_TOTAL.” If the number of people who were analyzed was presented, this was tagged as “ANALYSIS_TOTAL”. If the number of people who completed the trial or a certain follow-up period was presented, this was tagged as “COMPLETION_TOTAL. If the number of people who were part of the trial without being more specific was presented, this was tagged as “GENERAL_TOTAL”. If the number of people who were assigned to an arm of the trial was presented, this was tagged as “ARM”. Lastly, if the number of patients who were assigned to an arm was presented in the context of how many patients experienced an event, this was tagged as “ARM_EVENT”. As a hypothetical example, in the sentence “50 of 200 people in the intervention arm and 20 of 203 people in the control arm experienced treatment-related toxicity”, 200 and 203 would be tagged as ARM_EVENT. If the abstract did not contain the aforementioned entities, the manuscript was added to the dataset without any tags.

Note that while the dataset and code use the all-caps names for the entities, we will use normal names due to readability for the rest of the paper.150 annotated examples were randomly assigned to an unseen test set. The remaining 846 examples were used to train and validate a named entity recognition model using a random 85:15 split into training and validation. The transformer model RoBERTa-base was trained using Adam as the optimizer.^10,11^ The detailed configuration file with all parameters used for training and validation is available from the code repository at https://github.com/windisch-paul/sample_size_extraction.

In addition to training a named entity recognition model, we also built a system of regular expressions and conditional statements for cleaning the entities, e.g., to turn numbers presented as words into integers or to remove the “n=“ from an entity that was extracted as “n=934”.

After training the model, inference was done on the unseen test set. For each entity, we assessed its agreement with the ground truth. If the same entity was extracted several times from the same trial, we used two different approaches: In the case of entities that are supposed to indicate the total number of people in the trial, such as randomized total, analysis total, completion total, or general total, we used the maximum number of each respective entity. In the case of entities where each entity only presents a part of the people in the trial, such as arm and arm event, we summed up all instances of the same entity. After assessing the performance of the individual entities, we combined them into different models for a final prediction of the ground truth (i.e., how many people underwent randomization) using different conditions. The different models were supposed to have different strengths and weaknesses to allow for different use cases. For the first model, the ordered model, we simply ordered the entities according to their performance and built a model that returns the best-performing entity that is present in a publication. If no entity is identified, no prediction is made. For the second model, the accurate entities model, we only chose the three best-performing entities and instructed the model to refrain from making a prediction if none of these entities is found. For the third model, the conditional mode, we only allowed the model to make a prediction if either the best-performing entity was found or the second and third best-performing entity were both found and within 10% agreement of each other.

Training, validation, and testing were performed in Python (version 3.11.5) using, among others, the pandas (version 2.1.0), spacy (version 3.7.4), and spacy-transformers (1.2.5) packages.

## Results

The performance of the different entities is presented in Table 1. The arms entity was most frequently identified (in 74.0% of trials in the test set), followed by the general total (65.3%) and the randomized total (42.0%). The best performance in terms of predicting the ground truth was demonstrated by the randomized total, the arms, and the general total, which all demonstrated a median absolute percentage error of 0.0% and mean absolute percentage errors of 2.4%, 11.1%, and 19.0%, respectively. Scatterplots depicting the agreement between the extracted entities and the ground truth are presented in Figure 1.

**Table 1.** Performance of different entities. The “Extracted from” column indicates the percentage of trials in which the respective entity was found. The remaining columns indicate the accuracy of the respective entity in predicting the ground truth, i.e., how many people were randomized.

|  | Extracted from - % | Mean absolute percentage error - % | Median absolute percentage error - % | Extracted entities within 1% from ground truth - % | Extracted entities within 10% from ground truth - % |
| --- | --- | --- | --- | --- | --- |
| **Randomized total** | 42.0 | 2.4 | 0.0 | 95.2 | 96.8 |
| **Analysis total** | 12.7 | 38.0 | 1.3 | 47.4 | 89.5 |
| **Completion total** | 20.0 | 32.6 | 6.7 | 16.7 | 66.7 |
| **General total** | 65.3 | 19.0 | 0.0 | 72.5 | 85.7 |
| **Arms** | 74.0 | 11.1 | 0.0 | 72.9 | 81.9 |
| **Arm events** | 39.3 | 61.5 | 6.3 | 28.8 | 52.4 |

**Figure 1.**
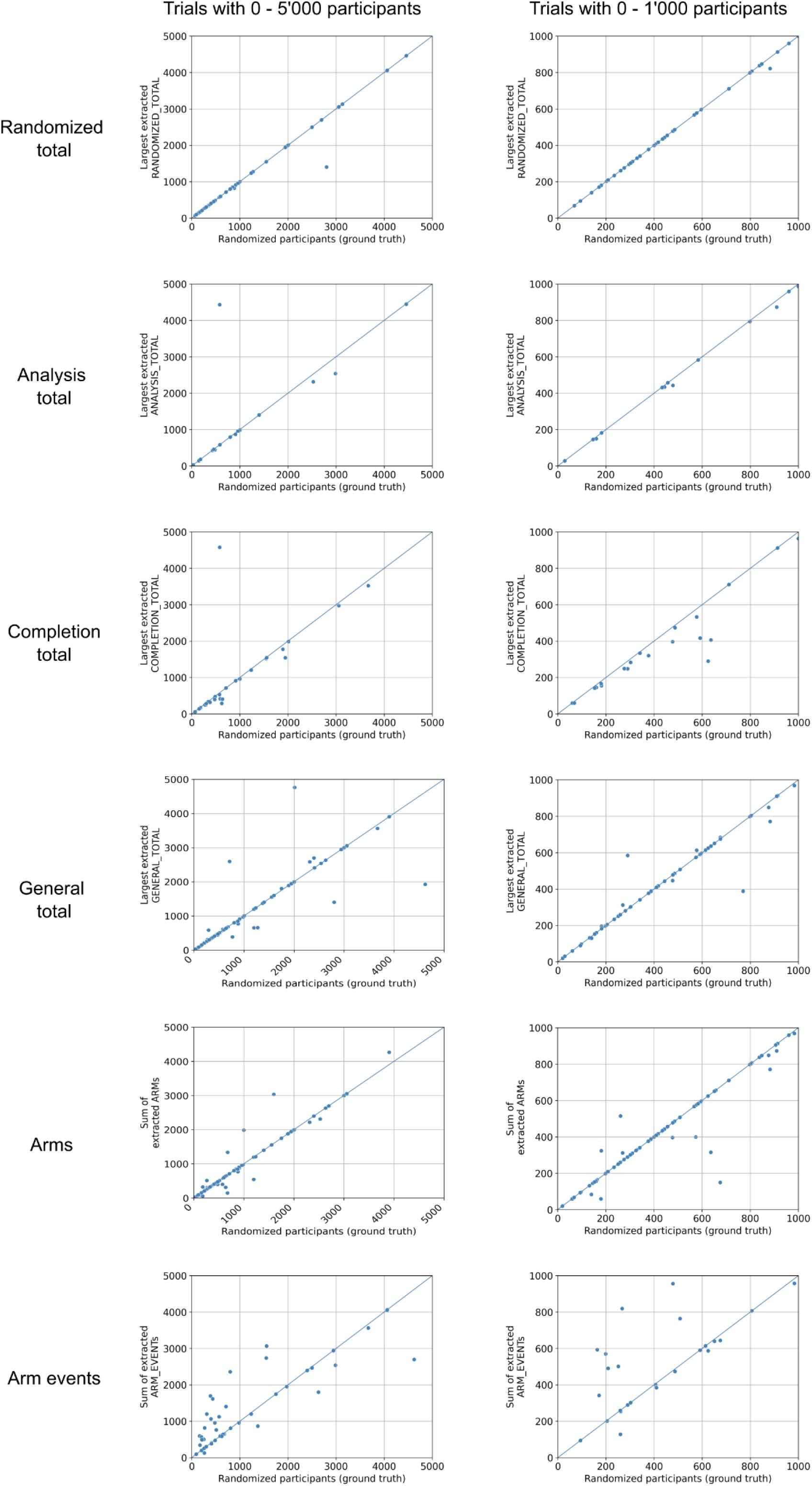
Scatterplots of different extracted entities. Each dot represents a trial with the ground truth, i.e., the number of participants who were randomized being its x-coordinate. The y-coordinate is the respective entity. For the randomized total, analysis total, completion total, and general total entities, the largest respective number was used in case multiple numbers were extracted from the same trial. In case of the arm and arm event entities, all extracted numbers from the same entity for a trial were summed up. Plots on the left show trials with 0 to 5’000 participants, while plots on the right show only trials with 0 - 1’000 participants to allow for a better assessment of the performance in the range that most trials fall into. The diagonal lines indicate perfect predictions.

The performance of the three models is depicted in Table 2 and Figure 2. While the conditional model exhibited the best performance and made predictions that were within 1% of the ground truth in 92.8% of cases and within 10% in 96.9% of cases, it could only make predictions on 64.7% of all the trials in the test set due to the strict conditions regarding entities that need to be present for making predictions.

**Table 2.** Performance of different models. The “Predicted in” column indicates the percentage of trials for which a prediction could be made. The remaining columns indicate the accuracy of the respective model in predicting the ground truth, i.e., how many people were randomized.

|  | Predicted in - % | Mean absolute percentage error - % | Median absolute percentage error - % | Extracted entities within 1% from ground truth - % | Extracted entities within 10% from ground truth - % |
| --- | --- | --- | --- | --- | --- |
| **Ordered model** | 98.0 | 13.7 | 0.0 | 78.9 | 86.4 |
| **Accurate entities model** | 96.0 | 7.9 | 0.0 | 80.6 | 88.2 |
| **Conditional model** | 64.7 | 1.82 | 0.0 | 92.8 | 96.9 |

**Figure 2.**
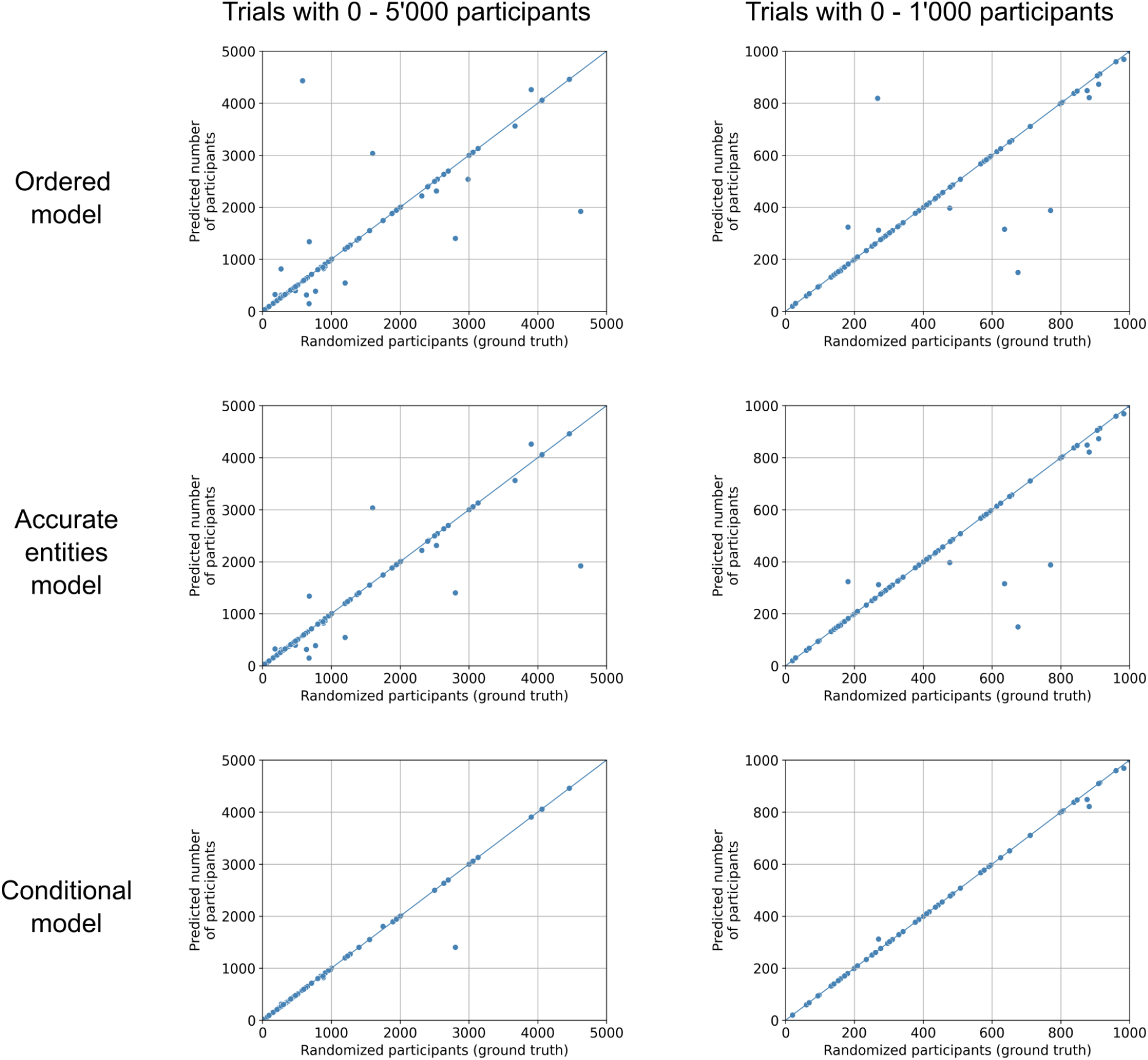
Scatterplots of different models. Each dot represents a trial with the ground truth, i.e., the number of participants who were randomized being its x-coordinate. The y-coordinate is the prediction by the model. Plots on the left show trials with 0 to 5’000 participants, while plots on the right show only trials with 0 - 1’000 participants to allow for a better assessment of the performance in the range that most trials fall into. The diagonal lines indicate perfect predictions.

On the other end of the spectrum, the ordered model was able to make a prediction on 98.0% of trials in the test set. However, these predictions were less accurate, with 78.9% being within 1% of the ground truth and 86.4% within 10%.

The accurate entities model represents a compromise. It was able to make a prediction on 96.0% of trials, with 80.6% of its predictions being within 1% of the ground truth and 88.2% within 10%.

## Discussion

This study presents a named entity recognition model that can detect six different entities in order to predict the sample size from the abstract of a randomized controlled trial (RCT). The entities can be combined in different ways to obtain models with different characteristics.

The fact that the randomized total entity demonstrated the best performance is unsurprising, considering that the number of people who were randomized was the ground truth and that sentences that explicitly state how many people were randomized should be very indicative of this. The completion and analysis total entities showed larger discrepancies. In the case of the completion total, the scatterplot suggests that this number is often smaller than the ground truth, which makes sense as not every patient who is randomized completes a trial. The same discrepancy can be seen with the analysis total, as not every trial reports its results by the intention-to-treat (which should be equal to the number of people who were randomized) but rather conducts a per-protocol analysis for which the number of included patients is often smaller. For the general total entity, there were also outliers where the extracted entity was larger than the ground truth. This happened, for example, when a trial stated that “X patients were included”, while the number X was actually the number of patients who were screened. In the case of the arm and arm event entities, the arm entity performs better and arm events produce more outliers where the entity was actually larger than the ground truth. This can happen due to multiple arm events being present. As an example, in a two-arm trial, the results might first report the results for the primary endpoint, then the rate of grade 2 toxicities and the rate of grade 3 toxicities. If, in each of those sentences, the number of patients in the arm is reported, all of those entities are extracted, and the sum of those entities is obviously larger than the actual number of patients in the trial. A possibility to improve this could be to train a model to predict how many arms a trial has and to only add up as many arm entities as there are arms in the trial.

The three models that were presented have different strengths and weaknesses that can support different use cases. The conditional model creates predictions that are highly accurate, but these predictions can only be made for around two-thirds of publications. A model like this could be used in a workflow where a human annotator manually annotates the other publications. The ordered and the accurate entities models could be used in a workflow where either a human annotator is not feasible, but accuracy is not as critical or where a human annotator manually annotates all trials and the tool pre-completes the annotations so that the annotator can accomplish his tasks with fewer clicks and, in turn, faster.^12^ In addition, one could also use different models and create an ensemble method. If all models agree, this indicates a high confidence level, and a manual review can be skipped, while disagreements warrant a closer look.

Comparing our models’ performances to previously published research is difficult. Both Lin and colleagues, as well as Marshall et al., in the publication proposing Trialstreamer report precision and recall and not continuous measures of performance compared to the ground truth.^5,9^ This makes sense for them, as their approach is to extract the sample size if it is explicitly mentioned in the paper. If the correct number is extracted, it is a true positive. If no sample size is present and the model does not extract a number, it is a true negative. Our approach is different in that we also predict the sample size even if it cannot be extracted directly, e.g., by extracting the number of patients in each arm and adding them up to allow for retrieving a sample size for more publications. Also, for some use cases, it might be acceptable to just get a rough estimate of how many patients were randomized, e.g., by using the number of patients who were analyzed or who completed the trial. However, the previous research results already supported the feasibility of this task in general, with a recall and precision of 0.79 and 0.88 presented by Marshall et al. and similar results presented by Lin and colleagues.

Our study is limited by the fact that we only used trials from seven journals for training and testing. While these are journals that publish many practice-changing randomized controlled trials, we can’t assess the model’s ability to generalize to trials from other journals, especially those that use unstructured abstracts. The strengths of this study include the use of a dedicated unseen test set and the high degree of reproducibility as all code and annotated data are shared in a public repository. The named entity recognition model returns entities that can be tailored by other researchers to make predictions based on the characteristics of the task that needs to be completed.

As an outlook, one might use larger language models for the same task in the future to see if improvements in performance can be obtained. Alternatively, one could try to link trial publications to their respective entry on clinicaltrials.gov and use the information there if the sample size cannot be inferred from the abstract. As a general observation, more strict enforcement of guidelines such as CONSORT could greatly improve the performance of text-mining efforts in evidence-based medicine.^13,14^ To enable readers to judge the model’s performance, a sample-size filter based on the model presented herein can be tested at https://www.scantrials.com/.

In conclusion, training a named entity recognition model to predict the sample size from randomized controlled trials is feasible, not only if the sample size is explicitly mentioned but also if the sample size can be calculated, e.g., by adding up the number of patients in each arm. Being able to extract the sample size automatically could support various meta-research efforts.

## Data Availability

All data and code used to obtain this study's results have been uploaded to .

https://github.com/windisch-paul/sample_size_extraction

